# Causal effects of serum levels of n-3 or n-6 polyunsaturated fatty acids on coronary artery disease: Mendelian randomization study

**DOI:** 10.1101/2020.10.18.20214791

**Authors:** Sehoon Park, Soojin Lee, Yaerim Kim, Yeonhee Lee, Min Woo Kang, Kwangsoo Kim, Yong Chul Kim, Seung Seok Han, Hajeong Lee, Jung Pyo Lee, Kwon Wook Joo, Chun Soo Lim, Yon Su Kim, Dong Ki Kim

## Abstract

**Background:** Additional studies on the causal effects of 3-n and 6-n polyunsaturated fatty acids (PUFAs) on the risk of coronary artery disease (CAD) are warranted.

**Methods:** This Mendelian randomization (MR) study utilized a genetic instrument developed from previous genome-wide association studies for various serum 3-n and 6-n PUFA levels. First, we calculated the allele scores for genetic predisposition of PUFAs in individuals of European ancestry in the UK Biobank data (N=337,129). The allele score-based MR was obtained by regressing the allele scores to CAD risks. Second, summary-level MR was performed with the CARDIoGRAMplusC4D data for CAD (N=184,305). The inverse variance-weighted or Wald ratio method was the main analysis for the summary-level MR, and when multiple single nucleotide polymorphisms were utilized (e.g., linoleic acid), MR-Egger and weighted median methods were implemented as sensitivity analyses.

**Results:** Higher genetically predicted eicosapentaenoic acid and dihomo-gamma-linolenic acid levels were significantly associated with a lower risk of CAD both in the allele-score-based and summary-level MR analyses. Higher allele scores for linoleic acid level were significantly associated with lower CAD risks, and in the summary-level MR, the causal estimates by the MR-Egger and weighted median methods also indicated that higher linoleic acid levels cause a lower risk of CAD. Arachidonic acid was the 6-n PUFA that showed significant causal estimates for a higher risk of CAD. Higher docosapentaenoic acid and adrenic acid levels showed inconsistent findings in the MR analysis results.

**Conclusions:** This study supports the causal effects of certain 3-n and 6-n PUFA types on the risk of CAD.

## Introduction

Coronary artery disease (CAD) is a comorbidity that critically affects patient prognosis and is associated with a substantial socioeconomic burden.^1^ The major goal of current medical interventions for metabolic disorders is to prevent CAD, but CAD is predicted to remain the primary cause of death worldwide along with the obesity issue and global aging trends. Thus, identifying protective or causative factors for CAD is an important health issue that may suggest preventive measures for CAD development.

As metabolic disorders are common predisposing factors for CAD,^2^ maintaining a healthy diet has been suggested to be an important lifestyle modification strategy for preventing CAD.^3^ Among the dietary components, dietary fat intake is one of the factors affecting CAD development, and controlling dyslipidemia is important for the primary prevention of CAD. Substituting saturated fats with unsaturated fats has been recommended in the guidelines and has shown benefits in reducing CAD risks in clinical studies.^4-8^ Among the unsaturated fats, polyunsaturated fatty acids (PUFAs), particularly n-3 and n-6 PUFAs, have been emphasized for their possible significant effect on the risk of cardiovascular diseases.^4,8^ However, the observed findings reported different CAD risks according to the PUFA subtypes^9^ and are inevitably prone to be affected by confounders or reverse causation. Thus, additional studies identifying the causal effects of various n-3 or n-6 PUFAs on the risk of CAD are warranted. However, although such evidence was presented by the GISSI trials for secondary prevention after the development of heart failure or coronary artery disease by n-3 PUFA supplementation,^10,11^ whether increasing PUFA intake may be helpful for primary prevention of CAD remains controversial due to heterogeneous intervention and the results of previous trials.^12,13^

Mendelian randomization (MR) is a useful tool for investigating causal effects from modifiable exposure to complex diseases.^14^ In MR, the exposure of interest is explained by a genetic instrument, and as one’s genotype is determined upon conception preceding the occurrence of confounders or diseases, MR can report causal estimates independent of confounding effects or reverse causation. MR has been implemented in the medical literature and has identified important causal factors for the risk of CAD or other chronic comorbidities.^15-18^

In this study, we aimed to investigate the causal effects of serum n-3 and n-6 PUFA levels on the risk of CAD by MR analysis testing the association between genetic predisposition for each PUFA type and the risk of CAD or myocardial infarction (MI). We performed both allele-score-based MR by individual-level data and MR based on summary-level data in different cohorts to replicate the findings. We hypothesized that certain n-3 or n-6 PUFAs would have causal effects on the risk of CAD.

## Methods

### Ethical considerations

The study was performed in accordance with the Declaration of Helsinki. The study was approved by the institutional review boards of Seoul National University Hospital (No. E-2070-048-1140) and the UK Biobank consortium (application No. 53799). As the study investigated anonymous databases or summary-level data, the requirement for informed consent was waived by the institutional review boards.

### Study setting

This study was an MR analysis including genetically explained exposures and outcomes from observational cohorts independent of the population where the genetic instrument was developed (i.e., two-sample MR). The study first utilized the UK Biobank data, which is the largest cohort to date with deep genotyping and collection of various clinicodemographic information.^19^ The UK Biobank data have been introduced to inspect the presence of possible confounder-associated variants in the genetic instrument and as the outcome data for allele-score based MR by the individual-level data. In addition, we performed summary-level MR with another independent observational genome-wide association study by the CARDIoGRAMplusC4D consortium. The analysis was performed to ask whether our findings can be replicated by another large-scale cohort by MR based on summary statistics.

### Genetic instrument

The study utilized two well-known genome-wide association meta-analysis results for the serum levels of specific types of n-3 and n-6 PUFAs of individuals of European ancestry.^20,21^ The study included genome-wide significant (P < 5×10^−8^) single nucleotide polymorphisms (SNPs) that were not in linkage disequilibrium (R^2^ < 0.1) with the PUFAs identified by the GWAS. The genetic instruments were repetitively utilized in the literature, including MR analysis, to study the causal effects of PUFA levels on various diseases and were identified to be on genes that are relevant to lipid metabolism.^22-24^ We utilized the genetic instruments for 3-n PUFAs [docosapentaenoic acid (3 SNPs), eicosapentaenoic acid (2 SNPs), and docosahexaenoic acid (1 SNP)] and for 6-n PUFAs [linoleic acid (3 SNPs), gamma-linolenic acid (2 SNPs), dihomo-gamma-linolenic acid (2 SNPs), adrenic acid (1 SNP), and arachidonic acid (2 SNPs)], and their summary statistics are presented in Supplemental Table 1.

The MR investigation requires three assumptions to be met to demonstrate causal effects.^14^ First, the relevance assumption means that the genetic instrument should be closely associated with the exposure of interest. As all included genetic variants reached genome-wide significance level association with each PUFA level and the variants were in functionally relevant genes for PUFA metabolism, the assumption was met. Second, the independence assumption indicates that the genetic instrument should not be associated with confounders or the absence of directional pleiotropy. The utilized genetic instrument for each PUFA phenotype included 1 to 3 SNPs with known functional relevance, which may decrease the possibility of heterogeneity. To additionally inspect that the independence assumption was met, we performed a separate genome-wide association study including the variants for the possible confounders hypertension, diabetes mellitus, and obesity in the UK Biobank data, adjusted for age, sex, and the first 10 principal components of the genetic information (Supplemental Table 2). However, no SNPs showed a genome-wide significant association with the confounders, so the original genetic variants remained in the implemented genetic instruments. The third assumption is the exclusion-restriction assumption, meaning that the causal effect should be only through the exposure of interest. Although this assumption is formally untestable, the previous observational findings suggest that serum n-3 and n-6 PUFA levels would be plausible factors to be tested for the causal effects on the risk of CAD.

### Allele-score based MR with individual-level data in the UK Biobank

The UK Biobank is a prospective population-based cohort of > 500,000 individuals aged 40-69 years from 2006 to 2010 in the United Kingdom. The details of the database have been published before.^19,25,26^ For genetic analysis, as the genetic instruments were developed in individuals of European ancestry, we included the UK Biobank data of individuals of white British ancestry. We excluded those who were outliers in terms of heterozygosity or missing rate, those with sex chromosome aneuploidy, and unrelated samples who were included in the genetic principal component calculations.^27^ The approach resulted in 337129 individuals included in the genetic analysis with the UK Biobank data.

In the allele-score-based MR, we assessed the risk of MI in the UK Biobank data, which was algorithmically defined by the UK Biobank and included death from MI and ST-segment elevated MI or non-ST-segment MI events based on hospital admission records and death registries. We included events through 29 February 2016, as complete hospital inpatient data were available until that date in all three regions of the nation: England, Scotland, and Wales.

We calculated allele scores for the exposures by multiplying the gene dosage matrix with the effect sizes of the genetic instrument by using PLINK 2.0 (version alpha 2.3).^28^ The associations between the genetic predisposition for each serum PUFA level represented by the allele scores and MI were investigated by logistic regression analysis, and age, sex, and the first 10 principal components were adjusted.^16^ We additionally performed a sensitivity analysis by adding phenotypical hypertension, diabetes mellitus, obesity, medication use for dyslipidemia, laboratory values of triglycerides, low-density lipoprotein, high-density lipoprotein and smoking history (none, ex-smoker, and current smoker) to the regression model.

The details regarding the data collection in the UK Biobank data are described in the Supplemental Methods. The regression analyses were performed using R (version 4.0.1, the R foundation), and two-sided P values <0.05 were considered significant.

### Summary-level MR with the CARDIoGRAMplusC4D data

Additional summary level-based MR was performed with the summary statistics provided by the CARDIoGRAMplusC4D study including participants mainly of European ancestry.^29^ We tested the causal estimates for the MI (43676 cases and 128199 controls) and CAD (60801 cases and 123504 controls) outcomes from the CARDIoGRAMplusC4D GWAS results. The fixed-effects inverse variance weighted method was the main MR method considering the small number of SNPs in each genetic instrument. When the number of SNPs was 3, for linoleic acid, additional sensitivity analysis by the penalized weighted median method,^30^ which gives valid causal estimates even when invalid instrument is present, and by MR-Egger regression with bootstrapped standard error,^31^ which yields pleiotropy-robust causal estimates, were performed. When a single SNP was included in a genetic instrument, the causal estimates were driven by the Wald ratio method. The above analyses were performed with the TwoSampleMR package in R.^32^

## Results

### Clinical characteristics of the UK Biobank data

The baseline characteristics of the UK Biobank participants of white British ancestry utilized for the genetic analysis are presented in Table 1. The median age was 58 years old, with 54% males and 46% females. The prevalence of hypertension and diabetes was 21% and 5%, respectively, with 18% of participants taking medication for dyslipidemia. The interquartile ranges for triglycerides, low-density lipoprotein, high-density lipoprotein, and estimated glomerular filtration rate values were within the reference ranges. The prevalent/incident MI outcome was identified in 12812 (4%) individuals.

**Table 1.** Baseline characteristics of the study population of the UK Biobank for the genetic analysis.

| Characteristics | Total (N = 337129) | Males (N = 156106) | Females (N = 181023) |
| --- | --- | --- | --- |
| Age (years) | 58 [51;63] | 59 [51;64] | 58 [51;63] |
| Body mass index | 26.7 [24.1;29.8] | 27.3 [25.0;30.0] | 26.1 [23.4;29.6] |
| Obesity (body mass index $\geq 30$ kg/m<sup>2</sup>) | 81022 (24.1%) | 39328 (25.3%) | 41694 (23.1%) |
| Smoking history |  |  |  |
| Non-smoker | 183636 (55%) | 76356 (49%) | 107280 (60%) |
| Ex-smoker | 118399 (35%) | 60835 (39%) | 57564 (32%) |
| Current-smoker | 33921 (10%) | 18360 (12%) | 15561 (9%) |
| Hypertension | 70018 (20.9%) | 38538 (24.9%) | 31480 (17.5%) |
| Systolic BP (mmHg) | 136.5 [125.0;149.5] | 139.5 [129.0;152.0] | 133.5 [121.5;147.5] |
| Diastolic BP (mmHg) | 82.0 [75.5;89.0] | 84.0 [77.5;90.5] | 80.0 [73.5;87.0] |
| Diabetes mellitus | 16178 (5%) | 10012 (6%) | 6166 (3%) |
| Hemoglobin A1c (mmol/L) | 35.1 [32.7;37.7] | 35.2 [32.7;37.9] | 35.1 [32.7;37.6] |
| Medications for dyslipidemia | 58531 (18%) | 35832 (23%) | 22699 (12.6%) |
| Triglycerides (mmol/L) | 1.5 [ 1.1; 2.2] | 1.7 [ 1.2; 2.5] | 1.3 [ 1.0; 1.9] |
| High-density lipoprotein (mmol/L) | 1.4 [ 1.2; 1.7] | 1.2 [ 1.1; 1.5] | 1.6 [ 1.3; 1.8] |
| Low-density lipoprotein (mmol/L) | 3.5 [ 3.0; 4.1] | 3.5 [ 2.9; 4.1] | 3.6 [ 3.0; 4.2] |
| Aspartate aminotransferase (U/L) | 20.2 [15.4;27.4] | 23.8 [18.4;31.8] | 17.5 [13.9;23.0] |
| Alanine aminotransferase (U/L) | 24.4 [21.0;28.8] | 26.1 [22.6;30.9] | 23.0 [20.0;26.8] |
| Creatinine (mmol/L) | 70.5 [61.6;81.0] | 80.0 [72.6;88.3] | 63.2 [57.1;70.0] |
| Estimated glomerular filtration rate<br>(mL/min/1.73 m<sup>2</sup>) | 92.5 [82.6;99.5] | 92.2 [82.6;99.3] | 92.9 [82.6;99.8] |
| Number of prevalent/incident MI cases | 12812 (4%) | 9878 (6%) | 2934 (2%) |

Categorical variables are presented as number (%) and continuous variables are presented as value [interquartile ranges].

### Allele-score based MR results

The causal estimates by the allele score-based MR are presented in Table 2. Among the 3-n PUFAs, genetically predicted eicosapentaenoic acid levels were significantly associated with lower odds for MI, while the allele score for docosapentaenoic acid was significantly associated with higher MI risks. Genetic predispositions for higher linoleic acid and dihomo-gamma-linolenic acid were significantly associated with lower risks of MI. On the other hand, genetically predicted gamma-linolenic acid and arachidonic acid levels were significantly associated with higher MI risks. Adrenic acid showed null causal estimates for the risk of MI. The above results were similarly reproduced even after we included additional phenotypical covariates in the regression analysis.

**Table 2.**
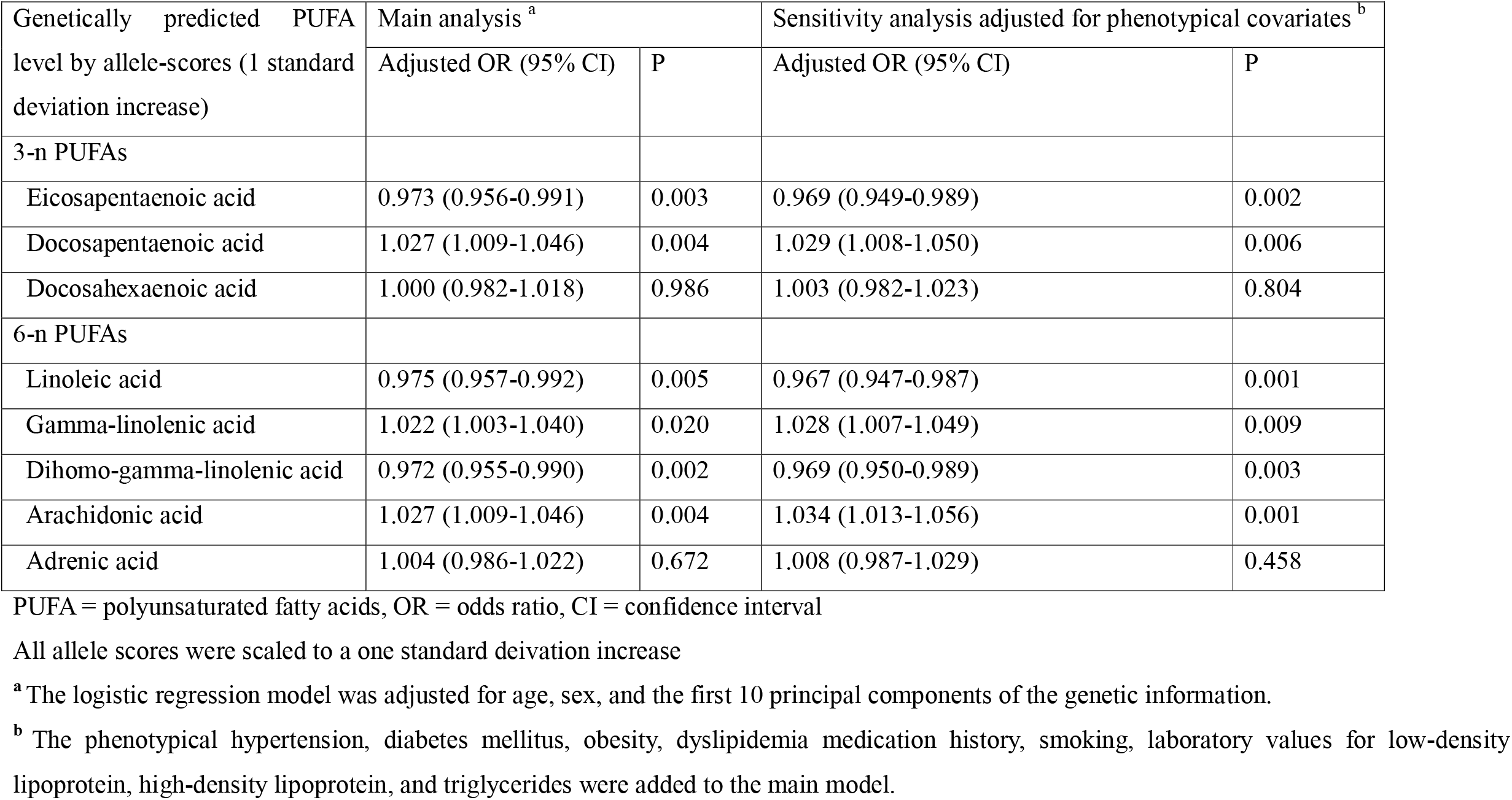
Allele-score based Mendelian randomization results in the UK Biobank data.

| Genetically predicted PUFA level by allele-scores (1 standard deviation increase) | Main analysis <sup>a</sup> |  | Sensitivity analysis adjusted for phenotypical covariates <sup>b</sup> |  |
| --- | --- | --- | --- | --- |
|  | Adjusted OR (95% CI) | P | Adjusted OR (95% CI) | P |
| 3-n PUFAs |  |  |  |  |
| Eicosapentaenoic acid | 0.973 (0.956-0.991) | 0.003 | 0.969 (0.949-0.989) | 0.002 |
| Docosapentaenoic acid | 1.027 (1.009-1.046) | 0.004 | 1.029 (1.008-1.050) | 0.006 |
| Docosahexaenoic acid | 1.000 (0.982-1.018) | 0.986 | 1.003 (0.982-1.023) | 0.804 |
| 6-n PUFAs |  |  |  |  |
| Linoleic acid | 0.975 (0.957-0.992) | 0.005 | 0.967 (0.947-0.987) | 0.001 |
| Gamma-linolenic acid | 1.022 (1.003-1.040) | 0.020 | 1.028 (1.007-1.049) | 0.009 |
| Dihomo-gamma-linolenic acid | 0.972 (0.955-0.990) | 0.002 | 0.969 (0.950-0.989) | 0.003 |
| Arachidonic acid | 1.027 (1.009-1.046) | 0.004 | 1.034 (1.013-1.056) | 0.001 |
| Adrenic acid | 1.004 (0.986-1.022) | 0.672 | 1.008 (0.987-1.029) | 0.458 |

PUFA = polyunsaturated fatty acids, OR = odds ratio, CI = confidence interval
All allele scores were scaled to a one standard deviation increase
<sup>a</sup> The logistic regression model was adjusted for age, sex, and the first 10 principal components of the genetic information.
<sup>b</sup> The phenotypical hypertension, diabetes mellitus, obesity, dyslipidemia medication history, smoking, laboratory values for low-density lipoprotein, high-density lipoprotein, and triglycerides were added to the main model.

**Table 3.** Summary-level Mendelian randomization results with the CARDIoGRAMplusC4D data.

| Genetically predicted PUFA level by allele scores | For coronary artery disease |  | For myocardial infarction |  |
| --- | --- | --- | --- | --- |
|  | OR (95% CI) | P | OR (95% CI) | P |
| 3-n PUFAs |  |  |  |  |
| Eicosapentaenoic acid | 0.781 (0.626-0.975) | 0.029 | 0.793 (0.537-1.172) | 0.245 |
| Docosapentaenoic acid <sup>a</sup> | 1.215 (0.971-1.522) | 0.089 | 1.227 (0.954-1.578) | 0.110 |
| Docosahexaenoic acid | 1.000 (0.851-1.175) | 0.999 | 1.057 (0.883-1.264) | 0.548 |
| 6-n PUFAs |  |  |  |  |
| Linoleic acid | 0.987 (0.975-1.000) | 0.055 | 0.986 (0.972-1.000) | 0.053 |
| Penalised weighted median <sup>b</sup> | 0.986 (0.974-0.999) | 0.035 | 0.984 (0.970-0.999) | 0.033 |
| MR-Egger <sup>b</sup> | 0.979 (0.958-1.000) | 0.024 | 0.975 (0.951-0.999) | 0.022 |
| Gamma-linolenic acid | 2.541 (0.783-8.244) | 0.120 | 2.960 (0.790-11.097) | 0.107 |
| Dihomo-gamma-linolenic acid | 0.940 (0.897-0.985) | 0.010 | 0.932 (0.884-0.983) | 0.009 |
| Arachidonic acid | 1.012 (1.000-1.024) | 0.042 | 1.014 (1.001-1.027) | 0.037 |
| Adrenic acid <sup>a</sup> | 1.587 (1.054-2.391) | 0.027 | 1.700 (1.073-2.693) | 0.024 |

PUFA = polyunsaturated fatty acid, OR = odds ratio, CI = confidence interval
<sup>a</sup> The causal estimates were driven by the Wald ratio method. Otherwise, the fixed effects inverse variance weighted method was implemented.
<sup>b</sup> Additional sensitivity analyses were performed as 3 SNPs were included in the genetic instrument for linoleic acid.

### Summary-level MR results

When the analysis was replicated with the CARDIoGRAMplusC4D data, the genetic predisposition for higher serum eicosapentaenoic acid levels was significantly associated with a lower risk of CAD. For docosapentaenoic acid, the direction of the ORs was consistent with the above allele score-based MR; however, the causal estimates did not reach the statistically significant level. For 6-n PUFAs, higher levels of genetically predicted linoleic acid were marginally associated with a lower risk of CAD or MI by the inverse variance-weighted method. When additional pleiotropy-robust methods were implemented, both the penalized weighted median and the MR-Egger regression results indicated that genetic predisposition for higher linoleic acid levels was significantly associated with lower risk of both CAD and MI. The causal estimates were similar to the findings in the UK Biobank data for dihomo-gamma-linolenic acid and arachidonic acid, as genetically explained dihomo-gamma-linolenic acid was significantly associated with lower CAD and MI risks, while arachidonic acid was causally linked to higher risks of CAD and MI. Gamma-linoleic acid, in which the causal estimates were significant in the allele score-based MR, showed marginally significant causal estimates with notably high odds ratios in the summary-level MR, whereas adrenic acid, which showed null findings in the allele score-based MR, showed significant causal estimates for higher risks of both CAD and MI in the CARDIoGRAMplusC4D data.

## Discussion

This study, including MR investigation, identified that serum levels of certain 3-n or 6-n PUFAs causally affect the risk of CAD or MI. Higher genetically predicted eicosapentaenoic acid and dihomo-gamma-linolenic acid levels were significantly associated with a lower risk of CAD or MI. Linoleic acid was also considered to be protective against CAD or MI in our results. Arachidonic acid was the 6-n PUFA that showed significant causal estimates for a higher risk of CAD or MI. Higher docosapentaenoic acid and adrenic acid may also be causative for CAD development, but the causal estimates were inconsistent or marginal in our investigations.

The clinical significance of 3-n and 6-n PUFAs has been debated. Several observational findings and meta-analyses reported the possible benefits of 3-n or 6-n PUFAs on the risk of cardiovascular disease.^4,8^ In addition, studies focusing on mechanisms of the protective effect of 3-n and 6-n PUFAs reported that PUFAs are related to atherogenesis, thrombotic activity, and inflammation.^33,34^ However, there were also contradictory reports addressing the absence of an effect of 3-n or 6-n PUFAs on CAD.^12^ That observational findings are inevitable for effects from coexisting confounders and for the potential issue of reverse causation limited the causal interpretation of the previous observational studies. Moreover, as the effects from specific PUFA types may vary, a clinical trial, which would reveal the causality of a PUFA on CAD, with a strict dietary modification for a single PUFA is difficult to perform, resulting in heterogeneous findings from previous trials.^12^ Thus, whether higher serum PUFA levels or levels of a specific PUFA can be effective for primary prevention of CAD has remained unanswered.

We performed this study to investigate the causal effects of various PUFA types on the risk of CAD or MI by implementing MR analysis. MR has a particular strength in that the method can reveal causal estimates from a modifiable exposure to complex diseases.^14^ The approach has now been widely introduced in the current medical literature and has reported causal effects from various exposures on complex diseases, which is difficult with randomized clinical trials. One’s genetic information is determined before the occurrence of any confounding factors or outcomes and is thus minimally biased by the effects from other clinical factors. In our MR analysis, we made efforts to attain the three key assumptions for an MR to demonstrate the causal effects. As the results were consistent for certain PUFA types, our study supports that serum PUFA levels causally affect the risk of CAD and MI. Moreover, our study reported that not all PUFA types uniformly reduce or increase the risk of CAD,^35^ and specific 3-n or 6-n PUFAs showed different causal directions. Thus, the findings may guide future trials that may prioritize specific PUFA types as interventional targets. Among the most abundant 3-n PUFAs, our MR findings indicated that higher serum eicosapentaenoic acid may causally decrease the risk of CAD, while the results for docosapentaenoic acid were inconsistent. A previous report also suggested that eicosapentaenoic acid may have a greater protective effect against CAD than docosapentaenoic acid. Additionally, supplementation with eicosapentaenoic acid has been reported to be preventive for CAD in hypercholesterolemic patients.^36^ Thus, eicosapentaenoic acid may be the prioritized 3-n PUFA as a supplemental target for the primary prevention of CAD. Regarding 6-n PUFAs, linoleic acid has been repeatedly reported for its importance in the risk of CAD, and experimental findings support its benefits on the cardiovascular system.^4,9,37^ The allele score-based MR and the pleiotropy-robust weighted median or MR-Egger analyses yielded significant causal estimates of linoleic acid on lower risk of CAD, which is consistent with the previous observational findings. A previous MR study reported null effects of linoleic acid on ischemic heart disease by utilizing different outcome summary statistics;^24^ however, as genetic instruments explain only a portion of the variance of an exposure, the results warranted additional validation. As genetic predisposition for higher linoleic acid was significantly associated with CAD or MI risks both in the CARDIoGRAMplusC4D and the UK Biobank data, the causal effects from linoleic acid on CAD would be interpreted to be present, supported by previous observational findings. In addition, for dihomo-gamma-linolenic acid, previous clinical and experimental studies reported that high levels of dihomo-gamma-linolenic acid may be protective against cardiovascular diseases.^35,38^ That an increased arachidonic acid level was causative for increased CAD or MI risks was also in concordance with previous observations reporting its proinflammatory role,^39,40^ and our study suggests the causality of its effect on the primary development of CAD. On the other hand, increased adrenic acid or gamma-linolenic acid serum levels were suspected to cause CAD or MI in the MR results;^41^ however, as the results were inconsistent in the replicative investigation, a future study is necessary to confirm their significance.

Some of the MR analysis results need additional explanation. First, the statistical power of an MR investigation generally increases by utilizing a large number of genetic variants to explain the exposure of interest.^42^ However, the currently available genetic instruments for each PUFA type included 1 to 3 SNPs, which may have low statistical power to capture the causal effects from genetic predispositions for PUFA levels. Additional investigations including a larger number of independent variants to genetically explain the serum PUFA levels may clearly distinguish the risk of CAD by an MR investigation. Next, as the actual size of an effect for relevant clinical intervention is different from the causal estimates in MR, the current MR results are qualitative information on the presence of causal effects from certain serum PUFA levels on CAD.^43,44^ Namely, one genetically predicted serum PUFA showing a larger effect size than another does not mean that the first serum PUFA would have larger clinical effects than the second. That the prevalence of CAD or MI was different between the UK Biobank and the CARDIoGRAMplusC4D data also explains the considerable differences in the sizes of the causal estimates by the two separate MR analyses. Additional clinical trials targeting different 3-n or 6-n PUFA subtypes are warranted to confirm whether the suggested causal effects from PUFAs on CAD development can be modified through interventions for PUFAs and to identify the clinical magnitude of the effects.

There are several limitations and unanswered questions in this study. First, as stated above, although this study suggested the causal effects of certain serum PUFA levels on CAD development, whether an effective intervention of modifying serum PUFA levels can actually be helpful for primary prevention of CAD should be answered by a future clinical trial. Second, additional experimental study is warranted to reveal the mechanism of different effects of certain serum PUFA levels on CAD risks. Third, as MR is weak for detecting nonlinear effects and as quantitative interpretation of our results is limited, the extent to which a serum PUFA level is beneficial or harmful cannot be answered by this study. Last, the study was mainly based on individuals of European ancestry due to data availability, so our results cannot be generalized to other ethnic populations.

In conclusion, this study supports the causal effect of certain 3-n and 6-n PUFAs on the risk of CAD or MI. A clinical trial targeting specific 3-n or 6-n PUFAs is warranted to reveal possible beneficial dietary interventions for the primary prevention of CAD.

## Data Availability

This study used the data from the UK Biobank consortium, and the approach was approved by the organization (application number: 53799). The summary statistics for the two-sample MR have been provided by the GIANT and the CKDGen consortia.

## Acknowledgements

The study was based on the data provided by the UK Biobank consortium (application No. 53799). We thank the investigators of the previous studies who provided valuable genetic summary statistics for this study.

## Funding

This work was supported by the Industrial Strategic Technology Development Program - Development of bio-core technology (10077474, Development of early diagnosis technology for acute/chronic renal failure) funded by the Ministry of Trade, Industry & Energy (MOTIE, Korea) and a grant from SNU R&DB Foundation (800-20190571). The study was performed independently by the authors.

## Conflicts of interest

None.

## Figure Legend

**Figure 1.**
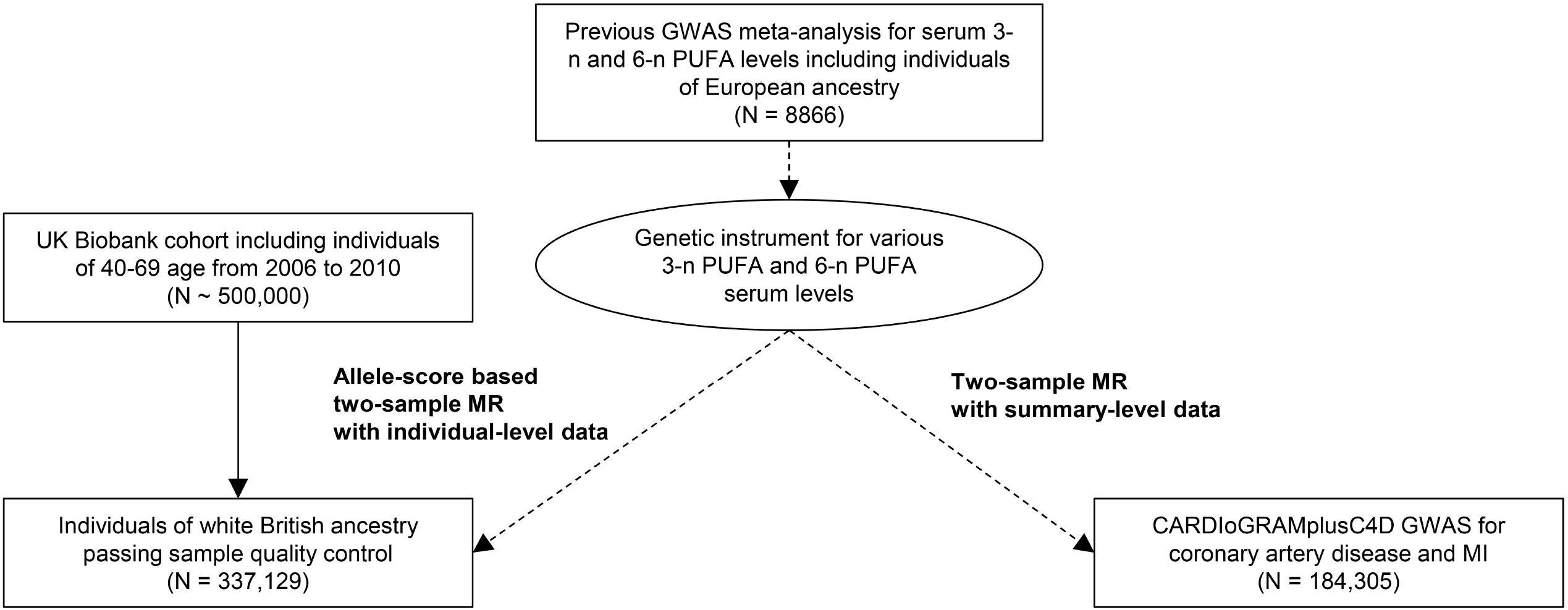
Study flow diagram. The study consisted of two parts; a two-sample MR analysis based on summary-level data with the CARDIoGRAMplusC4D data and an allele-score two-sample MR analysis based on individual-level data the UK Biobank data. PUFA = polyunsaturated fatty acid.

## Notes

### Competing Interest Statement

The authors have declared no competing interest.

### Clinical Trial

N/A

### Funding Statement

This work was supported by a grant from SNU R&DB Foundation (800-20190571) of the Seoul National University, Republic of Korea. The funders had no role in study design, data collection and analysis, decision to publish, or preparation of the manuscript.

### Author Declarations

The study was approved by the institutional review boards of Seoul National University Hospital (No. E-2070-048-1140) and the UK Biobank consortium (application No. 53799).

