## Supplemental Materials for "Causal effects of serum levels of n-3 or n-6 polyunsaturated fatty acids on coronary artery disease: Mendelian randomization study"

**Supplemental Table 1.** Summary statistics of the genetic instruments.

| Phenotype | SNP | Effect allele | Other allele | Effect allele frequency | Beta | Standard error |
| --- | --- | --- | --- | --- | --- | --- |
| Eicosapentaenoic acid | rs3798713 | C | G | 0.43 | 0.035 | 0.005 |
|  | rs174538 | A | G | 0.72 | 0.083 | 0.005 |
| Docosapentaenoic acid | rs780094 | T | C | 0.41 | 0.017 | 0.003 |
|  | rs3734398 | T | C | 0.43 | 0.04 | 0.003 |
|  | rs174547 | T | C | 0.67 | 0.075 | 0.003 |
| Docosahexaenoic acid | rs2236212 | C | G | 0.57 | 0.113 | 0.014 |
| Linoleic Acid | rs10740118 | G | C | 0.56 | 0.2484 | 0.0431 |
|  | rs174547 | C | T | 0.32 | 1.4737 | 0.0417 |
|  | rs16966952 | A | G | 0.31 | 0.3512 | 0.0439 |
| Gamma-linolenic acid | rs174547 | T | C | 0.67 | 0.0156 | 0.0009 |
|  | rs16966952 | G | A | 0.69 | 0.0061 | 0.0009 |
| Dihomo-gamma-linolenic acid | rs174547 | C | T | 0.33 | 0.355 | 0.0136 |
|  | rs16966952 | G | A | 0.69 | 0.22 | 0.013 |
| Arachidonic acid | rs174547 | T | C | 0.68 | 1.6909 | 0.0253 |
|  | rs16966952 | G | A | 0.69 | 0.1989 | 0.0314 |
| Adrenic acid | rs174547 | T | C | 0.67 | 0.0483 | 0.0019 |

**Supplemental Table 2.** Association between the genetic instruments and the phenotypical diabetes, hypertension, and obesity in the UK Biobank cohort.

| SNP | effect allele | Diabetes mellitus | | | Hypertension | | | Obesity | | | Associated PUFA phenotype |
| --- | --- | --- | --- | --- | --- | --- | --- | --- | --- | --- | --- |
|  |  | OR | Standard error | P value | OR | Standard error | P value | OR | Standard error | P value |  |
| rs10740118 | C | 0.984 | 0.012 | 0.163 | 0.976 | 0.006 | 0.000 | 0.974 | 0.006 | 5.16E-06 | Linoleic acid |
| rs174538 | A | 0.984 | 0.012 | 0.188 | 1.003 | 0.007 | 0.653 | 1.015 | 0.006 | 0.012 | Eicosaepentaenoic acid, Docosapentaenoic acid |
| rs174547 | C | 0.988 | 0.012 | 0.330 | 1.002 | 0.007 | 0.731 | 1.016 | 0.006 | 0.008 | Linoleic acid, Arachidonic acid, Dihomo-gamma-linolenic acid, gamma-linolenic acid, adrenic acid |
| rs16966952 | A | 0.994 | 0.013 | 0.605 | 0.994 | 0.007 | 0.390 | 0.988 | 0.006 | 0.046 | Linoleic acid, Arachidonic acid, Dihomo-gamma-linolenic acid, gamma-linolenic acid |
| rs780094 | T | 0.920 | 0.012 | 0.000 | 1.012 | 0.006 | 0.062 | 0.979 | 0.006 | 0.000 | Docosaepentaenoic acid |
| rs3734398 | C | 1.005 | 0.012 | 0.681 | 0.998 | 0.006 | 0.761 | 0.996 | 0.006 | 0.506 | Docosaepentaenoic acid |
| rs2236212 | C | 1.007 | 0.012 | 0.540 | 0.997 | 0.006 | 0.668 | 0.996 | 0.006 | 0.510 | Docosaehexaenoic acid |
| rs3798713 | C | 1.008 | 0.012 | 0.504 | 0.997 | 0.006 | 0.631 | 0.996 | 0.006 | 0.472 | Eicosaepentaenoic acid |

The genome-wide-association study results are adjusted for age, sex, and the first 20 genetic principal components. As no SNPs reached genome-wide significance level (5×10^-8^) with the three possible confounders, the utilized SNPs remained in the genetic instrument for further analyses.
